# PharmGScore scores of compound genetic variant burden for psychiatric treatment optimization

**DOI:** 10.1101/2023.06.27.23291888

**Authors:** Malgorzata Borczyk, Jacek Hajto, Marcin Piechota, Michal Korostynski

## Abstract

The acceptability of antidepressant drugs partly depends on genetic factors. The list of genes involved in antidepressant response, including Adverse Drug Reactions (ADRs) is broad and contains both drug-metabolizing enzymes (pharmacogenes) and genes involved in pharmacodynamics. Variants in pharmacogenes are traditionally reported in the form of star alleles and are partially annotated with known phenotypic consequences. As it is unfeasible to analyze all genotype-phenotype pairs, computational approaches remain the practical solution. A pharmacogenetic framework to predict responses to antidepressant drug treatment would provide great benefit to patients. In this study, we present a scoring system (PharmGScore) to assess both rare and common genetic variant burden across multiple genes. The PharmGScore is constructed by normalizing and aggregating existing, well-established computational variant predictors (CADD, Fathmm-xf, PROVEAN, Mutation Assessor). We show that this score effectively distinguishes no and decreased function from normal and increased function pharmacogenetic variants reported in PharmVar (PharmGScore AUC = 0.86). PharmGScore has improved performance when compared to its component scores (AUCs: CADD = 0.79; FATHMM-XF = 0.81; PROVEAN = 0.81; Mutation Assessor = 0.75). We then apply the PharmGScore to the 200k exome sequences of the UK Biobank (UKB). We report the overrepresentation of UKB participants with high (>50) gene PharmGScore for *CYP2C19* and *CYP2C9* and with high (>100) compound PharmGScore from nine pharmacogenes within a group with an antidepressant toxicity diagnostic code (T43.2). We then analyze all UKB participants that received any antidepressant toxicity or ADR diagnosis (n = 602). We indicate genes for which a higher burden may be associated with antidepressant toxicity or ADRs and confirm the known roles of *CYP2C19* and *CYP2D6* in this process. Finally, we show that patients who experienced ADRs to antidepressants in the therapeutic process or accidental poisoning with antidepressants have a higher PharmGScore composed of nine cytochrome P450 genes. Our study proposes a novel paradigm to assess the compound genetic variant burden associated with antidepressant response from exome sequencing data. This approach can be further applied to a user-defined set of genes to investigate other pharmacological traits.

## Introduction

Genome and exome sequencing are starting to become standard practices in the clinical world and show potential to improve both the understanding and the treatment of psychiatric patients (Borczyk et al. 2020.; Sanders et al. 2017). One prominent area of psychiatric genetics is the possibility of treatment personalisation. Personalized approaches to mental disorders have fundamental importance, as treatment resistance affects 20-60% of patients and there are not many new drugs in development (Howes, Thase, and Pillinger 2022). Depression is the most prevalent mental disorder, as it is the largest single contributor to health loss for humanity as a whole (World Health Organization. 2017). Pharmacogenomics studies the relationship between the variability in the genome sequence and drug responses (Lauschke and Ingelman-Sundberg 2019). One of the major areas for depression treatment personalization is in pharmacogenomics of antidepressant drugs as individual variability in antidepressant responses has a substantial genetic component (Iniesta et al. 2018; Tansey et al. 2013).

Variants in two known pharmacogenes, namely *CYP2D6* and *CYP2C19* have already been translated into effective clinical guidelines for antidepressant treatment (van Westrhenen et al. 2021). However, these guidelines only include known, relatively common, haplotypes of these genes, reported in the form of star alleles and their known phenotypes. This is an important limitation as 30-40% of interindividual variability in drug responses can be explained by rare genetic variants (Minor Allele Frequency -MAF < 1%) (Kozyra, Ingelman-Sundberg, and Lauschke 2017). As exome sequence data are becoming more and more available, the star allele approach needs updating to optimally incorporate patients’ genomic information for better treatment personalisation.

Another large caveat of current pharmacogenetic testing is that only a limited number of genes is included in the guidelines. It is known that antidepressant action is dependent on multiple genes, with ∼100 genes mentioned by the PharmGKB database (Whirl-Carrillo et al. 2021). These genes include both drug-metabolizing enzymes, as well as neurotransmitter receptors and drug transporters in the Central Nervous System (CNS). A computational approach that takes into account the combined effects of genetic variants across multiple genes would be beneficial. Another important point is that genes expressed in the CNS are typically very evolutionary constrained, therefore rare variants in these genes could be involved in determining antidepressant action (Karczewski et al. 2020). For example, a variant in the serotonin transporter *SERT* affecting an amino acid crucial in antidepressant binding could result in a drug being inactive (Andersen et al. 2010). Even if such variants are ultrarare or only exist in single individuals, an approach to identify these patients in an early phase of treatment could help to eliminate some of the difficulties leading to treatment resistance.

Attempts to build burden scores targeting pharmacogenes have been made previously (Zhou et al. 2019). The aim of our study was to improve this approach, creating the PharmGScore, a genetic burden score, based on the normalization of already established computational predictors for single variants like the CADD score (Rentzsch et al. 2019). PharmGScore allows for combining the burden of multiple pharmacogenetic variants into one value. Here we tested the performance of the PharmGScore using the PharmVar database and then applied it to the 200k Whole Exome Sequences (WES) available in the UK Biobank (UKB) database (Sudlow et al. 2015). The study then focused on the association of higher values of PharmGScore for the cytochrome P450 and other relevant genes with Adverse Drug Reaction (ADR) diagnoses to antidepressants. Overall, the presented approach is the first step toward an applicable computational pharmacogenetic framework.

## Materials and Methods

### Data and code availability

All the code used in this project is available in a GitHub repository (https://github.com/ippas/ifpan-pharmgscore-manuscript). The star allele definitions were downloaded from the Pharmacogene Variation Consortium (PharmVar v 5.1.6) at www.pharmvar.org (Gaedigk et al. 2021). The component scores of the PharmGScore are publicly available: CADD (Rentzsch et al. 2019), FathmmXF (Rogers et al. 2018), PROVEAN (Choi and Chan 2015) and MutationAssessor (Reva, Antipin, and Sander 2011). The UK Biobank data used in this research is available to approved researchers and has been accessed through the application 62979 “Impact of pharmacogenetic profiles on depression treatment outcomes”. The pre-computed PharmGScore for all possible single-nucleotide variants (SNV) in WES is available for download as tsv: https://pharmgscore.labpgx.com/ (Supplementary Table A)

### Data processing, phenotyping and variant score development

#### Software and environment

Analyses described below were performed with Hail (v. 0.2) - Python-based (v. 3.7) library for working with genomic data (https://hail.is/) and with R software environment (v. 4.2).

#### PharmGScore burden score construction

Protein-coding genes (19342 genes) along with their names and exon positions were downloaded from BioMart database (GRCh38.p13). In the score-building process we aimed to combine modern, machine-learning based scores for non-coding variants (e.g. CADD, FATHMM-XF) with scores specifically assessing deleteriousness of amino acid substitutions (e.g. PROVEAN, Mutation Assessor). For each group, we chose a pair of well-performing scores that exhibit closest-to-normal distributions of scores for pharmacogenetic variants (as shown by Zhou et al. 2019). CADD is a widely-used measure built from more than 60 genomics features with a machine-learning model. The model is trained on a binary distinction between simulated *de novo* variants and real human genetic variants. This way the training set for CADD is larger than the typical set of “known” damaging mutations (Rentzsch et al. 2019). FATHMM-XF is a supervised machine learning model, based on 27 annotations, trained to discriminate pathogenic mutations from the Human Gene Mutation Database and a subset of common, neutral, mutations from the 1000 genome project (Rogers et al. 2018). PROVEAN is an alignment-based score that compares homology levels of related protein sequences to predict the deleteriousness of mutations, while Mutation Assessor evaluates conservation of a given amino acid and has been validated on disease-causing polymorphisms (Choi and Chan 2015; Reva, Antipin, and Sander 2011). All possible single nucleotide substitutions variants within the range smaller than or equal to 50 bp from exons were annotated with the above-described four diverse scores: CADD, FATHMM-XF, PROVEAN and Mutation Assessor.

To combine the component scores into a single metric, we performed quantile normalization of each of the four scores to make the distribution the same as that defined by a target function. As this procedure was conducted separately for each gene the final PharmGScore is controlled for varying gene lengths. The target function is defined by: e^*x*^−e^−5^ for *x* ∈[−5.5] with the following properties: 1) possible range of the score from 0 to ∼150; 2) about a half of all variants with a score < 1 (based on the assumption that variants below the 50th percentile for each score should be neutral); 3) a widely spread range of scores (from 50 to 150) for the top 10% of most damaging variants (this spread allows for the score to distinguish moderately and highly deleterious variants). PharmGScore for each variant was defined as an average from non-missing normalized scores. For non-coding regions and synonymous variants only CADD and FathmmXF were used. In its basic form the PharmGScore scores individual variants, but it can be applied to groups of variants (e.g. within a star allele, gene or a set of genes) in a form of a sum of all individual variant scores. For star-allele and gene-level PharmGScore cut-off threshold of 50 (within the top 10% of most damaging variants) was used to determine the PharmGScore to be high - an equivalent to the PharmVar “no function” assignment.

#### Phenotyping and selection of UKB participants for analyses

Groups were phenotyped based on the hospital summary diagnoses and death registers available in the UKB. The first part of the study (i.e. results in Table 1) evaluated individual diagnostic codes associated with psychiatric drug toxicity/ADRs. To ensure sufficient statistical power only codes with more than 200 sequenced UKB European participants were included. The following International Classification of Diseases 10th Revision (ICD10) codes were studied in this part: T43.2 - poisoning by other and unspecisied antidepressants, T42.4 - poisoning by benzodiazepines, T40.2 - poisoning by other opioids, Y45.0 - opioids and related analgesics causing adverse effects in therapeutic use, T42.6 - poisoning by other antiepileptic and sedative-hypnotic drugs.

**Table 1.**
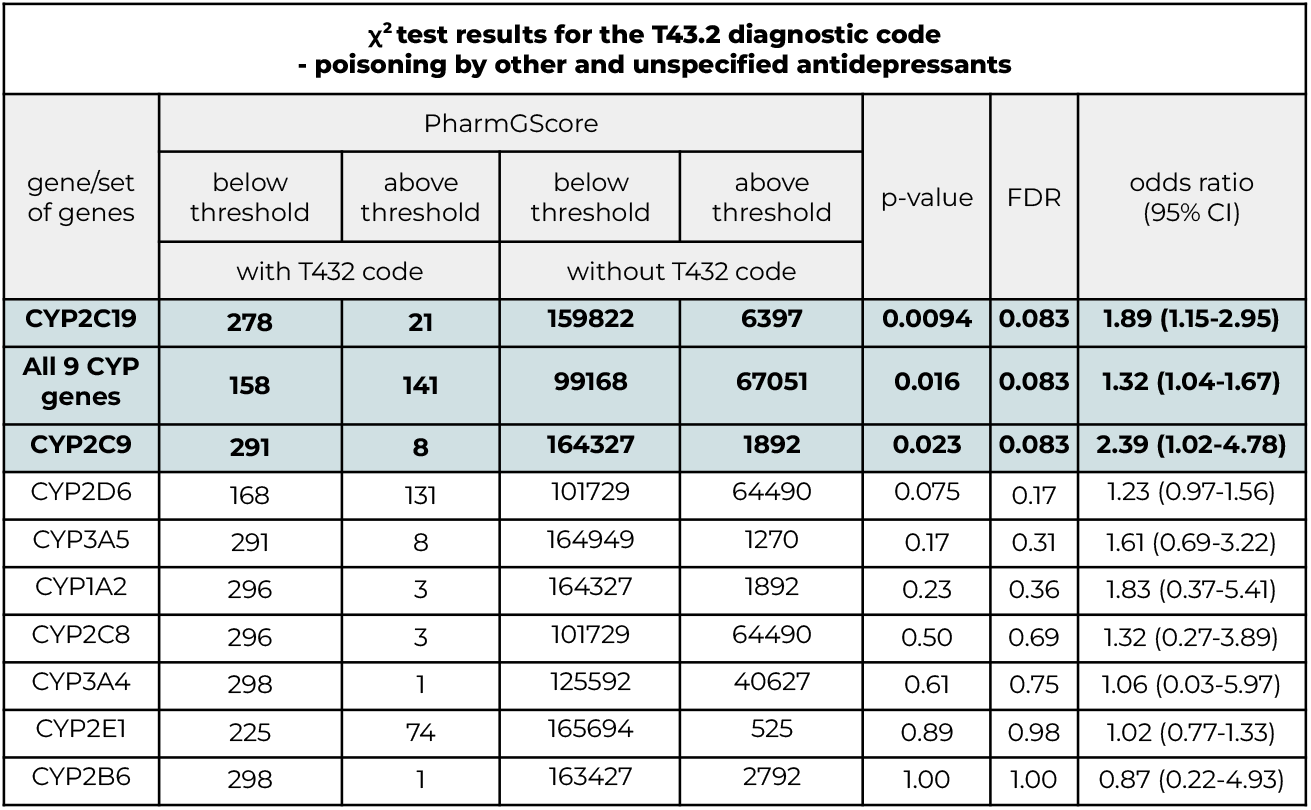
Chi-square test results for participants with high gene-level or compound PharmGScore versus those with low gene-level or compound PharmGScore for the T43.2 - poisoning by other and unspecified antidepressants diagnostic code. Results for the extended gene set and four other diagnostic codes are available in the supplementary table E.

The second part of the analyses (i.e. results in Figure 3, Tables 2 and 3) was performed with focus on all-cause antidepressant toxicity / ADRs. The main analyzed group, termed the ADR group (n = 602), consisted of UKB participants that were assigned any code related to antidepressant toxicity (T43.0-T43.2) or adverse effects (Y49.0 - Y49.2) in summary diagnoses (n = 592) or death register (n = 10). This group was further subdivided into the intentional subgroup and the accidental poisonings group. The inclusion criteria for the intentional poisonings were: antidepressant toxicity diagnostic code and a self-harm with drugs code at any time (any code from the X6 chapter). For the accidental subgroup participants with ADRs to antidepressants at therapeutic doses (Y49.0 - Y49.2) or with antidepressant poisoning (T43.0-T43.2) and no codes from the self-harm with drugs chapter (X6) were assigned. This means that participants with any history of intentional overdosing of any drugs were excluded from the accidental poisoning / ADR group. The ICD10 diagnostic code criteria for the control groups were as follows: all controls – all unrelated UKB samples from the 200k WES release not included in the ADR groups; depression and anxiety control group - samples from all controls that had any of the diagnostic codes describing depression and anxiety (F32.9, F33.9, F32.2, F32.1, F32.3, F33.1, F33.4, F25.1, F31.5, F33.8, F32.8, F31.3, F33.0, F31.6, F33.2, F41.9, F41.0, F41.1, F41.8, F60.6, F40.8, F40.9, F41.3); self-harm with drugs controls - participants from the all controls group that had any diagnostic codes from the self-harm with drugs chapter (X6).

**Table 2.**
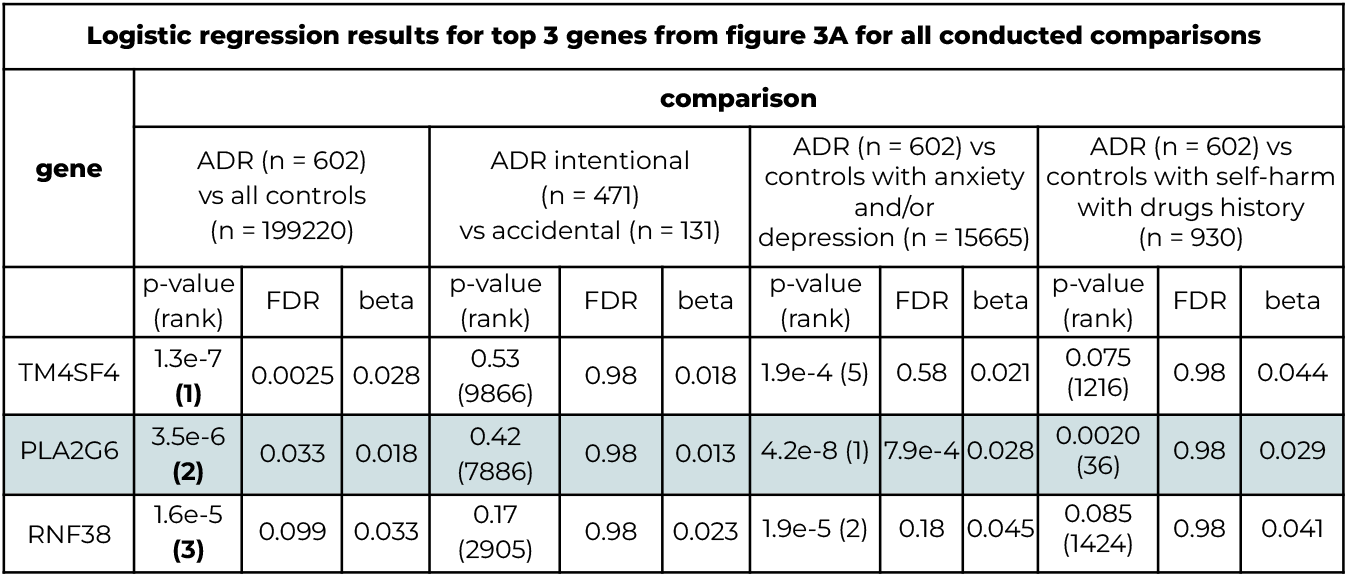
Results for top 3 genes from figure 3A for all performed comparisons.

| Logistic regression results for top 3 genes from figure 3A for all conducted comparisons |  |  |  |  |  |  |  |  |  |  |  |  |
| --- | --- | --- | --- | --- | --- | --- | --- | --- | --- | --- | --- | --- |
| gene | comparison |  |  |  |  |  |  |  |  |  |  |  |
|  | ADR (n = 602)<br>vs all controls<br>(n = 199220) |  |  | ADR intentional<br>(n = 471)<br>vs accidental (n = 131) |  |  | ADR (n = 602) vs<br>controls with anxiety<br>and/or<br>depression (n = 15665) |  |  | ADR (n = 602) vs<br>controls with self-harm<br>with drugs history<br>(n = 930) |  |  |
|  | p-value<br>(rank) | FDR | beta | p-value<br>(rank) | FDR | beta | p-value<br>(rank) | FDR | beta | p-value<br>(rank) | FDR | beta |
| TM4SF4 | 1.3e-7<br>(1) | 0.0025 | 0.028 | 0.53<br>(9866) | 0.98 | 0.018 | 1.9e-4 (5) | 0.58 | 0.021 | 0.075<br>(1216) | 0.98 | 0.044 |
| PLA2G6 | 3.5e-6<br>(2) | 0.033 | 0.018 | 0.42<br>(7886) | 0.98 | 0.013 | 4.2e-8 (1) | 7.9e-4 | 0.028 | 0.0020<br>(36) | 0.98 | 0.029 |
| RNF38 | 1.6e-5<br>(3) | 0.099 | 0.033 | 0.17<br>(2905) | 0.98 | 0.023 | 1.9e-5 (2) | 0.18 | 0.045 | 0.085<br>(1424) | 0.98 | 0.041 |

**Table 3.** Results for top 3 genes from figure 3B for all performed comparisons.

| Logistic regression results for top 3 genes from figure 3B for all conducted comparisons |  |  |  |  |  |  |  |  |  |  |  |  |
| --- | --- | --- | --- | --- | --- | --- | --- | --- | --- | --- | --- | --- |
| gene | comparison (for 90 selected genes) |  |  |  |  |  |  |  |  |  |  |  |
|  | ADR (n = 602)<br>vs all controls<br>(n = 199220) |  |  | ADR intentional<br>(n = 471)<br>vs accidental (n = 131) |  |  | ADR (n = 602) vs<br>controls with anxiety<br>and/or<br>depression (n = 15665) |  |  | ADR (n = 602) vs<br>controls with self-harm<br>with drugs history<br>(n = 930) |  |  |
|  | p-value<br>(rank) | FDR | beta | p-value<br>(rank) | FDR | beta | p-value<br>(rank) | FDR | beta | p-value<br>(rank) | FDR | beta |
| CYP2C19 | 0.0029<br>(1) | 0.27 | 0.0050 | 0.57<br>(48) | 0.96 | 0.0027 | 0.0079<br>(1) | 0.71 | 0.0045 | 0.026 (3) | 0.71 | 0.0052 |
| PPP2CB | 0.014 (2) | 0.61 | 0.025 | 0.88<br>(83) | 0.96 | 0.0088 | 0.037 (3) | 0.80 | 0.026 | 0.19 (18) | 0.77 | 0.052 |
| CYP2D6 | 0.030<br>(3) | 0.74 | 7.59e-4 | 0.39<br>(32) | 0.96 | -7.94e-4 | 0.029 (2) | 0.80 | 7.9e-4 | 0.013 (1) | 0.65 | 0.0011 |

#### Gene selection

Preselected sets of genes were used in a subset of the analyses. These included: 9 cytochrome P450 genes (9 genes: *CYP2C19, CYP2C9, CYP2D6, CYP3A5, CYP2B6, CYP1A2, CYP2C8, CYP3A4* and *CYP2E1*) and a list of other antidepressant-relevant genes (81 genes). The genes were selected from PharmGKB clinical annotations and literature. We included genes associated with efficacy, toxicity, metabolism, pharmacodynamic and pharmacokinetic pathways or ADRs to antidepressants (Whirl-Carrillo et al. 2021). Additionally, we added genes from two meta-analyses of pharmacogenetic studies of antidepressant responses, two large Genome-Wide Association Studies (GWAS) studies, and a targeted exome sequencing study (Crisafulli et al. 2011; C. Fabbri et al. 2018; Porcelli et al. 2011; Xu et al. 2020). Lists of included genes are available in the Supplementary Material.

### Statistical analyses of the PharmGScore and antidepressant responses

#### PharmGScore validation on PharmVar alleles

Results of this analysis are presented in Figure 1 and Supplementary Figures 1 and 2. We downloaded all star allele definition .vcf files from PharmVar (listed in Supplementary Table B). Each star allele (including sub star alleles) was considered a separate unit, thus individual single-nucleotide variants could have been included in more than one star allele. The star alleles with undefined phenotype and defined as ‘other’ were not considered (Gaedigk et al. 2021). We summed PharmGScores of all SNVs comprising each star allele to compute its final PharmGScore.

**Figure 1.**
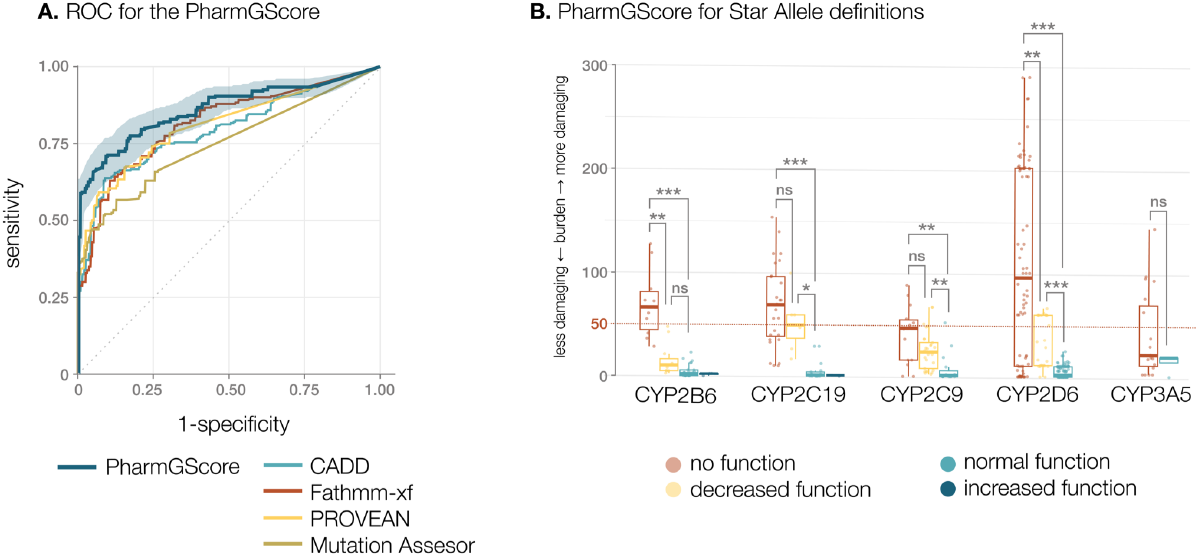
PharmGScore effectively predicts known phenotypes based on star alleles for selected genes. **A**. The Receiver-operator characteristics (ROC) curves for PharmGScore and its component scores. AUC of the ROCs: PharmGScore = 0.86, CADD = 0.79, FATHMM-XF = 0.81, PROVEAN = 0.81, Mutation Assessor = 0.75. For ROC normal and increased function star alleles were assigned a value of 0, while no function and decreased function alleles were assigned a value of 1. The 95% Confidence Interval (CI) for the PharmGScore is shaded with transparent blue. ROCs for the ability of the score to differentiate between individual phenotypes are presented in Supplementary Figure 2. **B**. Comparison of the PharmGScore for star alleles (summed from all SNVs) with known phenotypes for 5 cytochrome P450 (CYP) genes, most relevant to antidepressant metabolism and included in PharmVar (Whirl-Carrillo et al. 2021). The cut-off threshold of 50 (dotted line) was determined as the optimal threshold across selected genes. Differences between phenotypes for each gene were determined with one-way ANOVA and pairwise t-tests with Bonferroni correction (full statistical results are available in Supplementary Table C). Bonferroni corrected p-values: ns: not significant, *p < 0.05, **p < 0.01, ***p < 0.001

To assess PharmGScore performance, ROC-AUC (Area Under the Receiver operating characteristic Curve) of PharmGScore was compared with ROC-AUCs for its component scores. In the main assessment of the star allele PharmGScore we grouped the decreased and no function alleles together into one phenotype, and normal and increased function phenotypes into another. This grouping was chosen as Intermediate Metabolizers (with decreased function) of *CYP2C19* and *CYP2D6* genes have distinct clinical recommendations from normal metabolisers for several antidepressants (Abdullah-Koolmees et al. 2021). ROC-AUC analyses for each phenotype separately were additionally performed. A ROC-AUC assessment of PharmGScore performance depending on the distance from the exon that is used to include loci was done with the same grouping as the main evaluation. To compare average star allele PharmGScores (Figure 1B) one-way ANOVA and pairwise t-tests with Bonferroni correction were used.

### Annotation of the UK Biobank WES data with the PharmGScore

We performed variant and sample quality control of the 200k UKB WES as in a recent study of rare variants in the same dataset (Kingdom et al. 2022). We then annotated the remaining variants 200k UKB WES with the PharmGScore (Figure 2). First, related individuals were removed preferentially keeping participants that were assigned ADRs to antidepressants. We assumed a kinship cut-off 0.125 on a scale from 0 to 0.5, which removes second and closer degree relatives, similarly to a large UKB exome variant burden study (Karczewski et al. 2022). This left 199822 participants. Each protein-coding gene for each participant was then annotated with the sum of all single-variant PharmGScore to determine gene-level PharmGScores.

**Figure 2.**
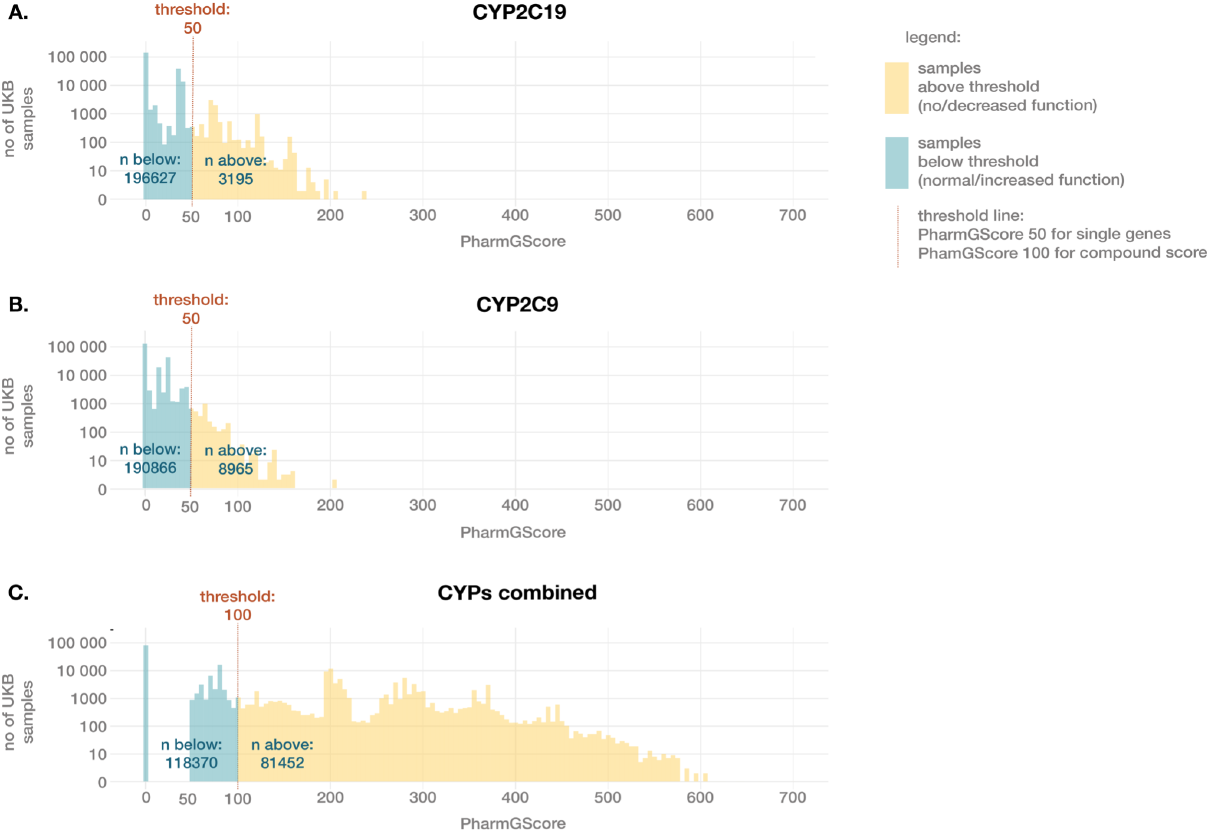
Distributions of the gene-level PharmGScore in 200k UK Biobank exomes for two example genes and a compound PharmGScore of a set of 9 cytochrome P450 genes. Here the gene-level PharmGScore distribution for 2 (A) *CYP2C19* and (B) *CYP2C9* and (C) a compound PharmGScore distribution for all 9 genes are shown as examples. These three cases were chosen for the figure as they had a significant overrepresentation of antidepressant poisoning diagnoses in the groups with high PharmGScore (see Table 1 and Supplementary Table E). The high PharmGScore threshold for individual genes is equal to 50, while for the compound score it is set to 100 - equivalent to two genes from the list above the single-gene threshold. For the compound score only genes with gene-level PharmGScore > 50 are included. The distribution of the PharmGScore for other analyzed CYP genes is presented in Supplementary Figure 3.

### Chi-square tests for 5 psychotropic drug toxicity diagnostic codes

For analysis overrepresentation of participants with high PharmGScore in selected diagnostic codes (T43.2, T4.24, T40.2, Y45.0, T42.6) (Table 1), the chi-square test was used if the smallest group was > 100 and Fisher exact tests were used for smaller groups. For the chi-square tests, the genetic ethnicity UKB field was used to filter only for European participants, which are the largest population in the database. This left 166518 samples. A selected test was additionally performed in the group of non-European participants (n = 33304).

### Single-gene logistic regression analyses

To compare variant burden between the main antidepressant ADR group and control groups (Figure 3 and Tables 2 and 3), logistic regression Wald’s test was applied. Here we included all UKB participants regardless of ethnicity and used precomputed UKB 17 genetic Principal Components (PC), age, sex and Townsend deprivation index as covariates. P-values are reported as nominal p-values and as corrected p-values with the Benjamini-Hochberg FDR method. This analysis was performed in two parts: for all protein-coding genes and for selected antidepressant-relevant genes.

**Figure 3.**
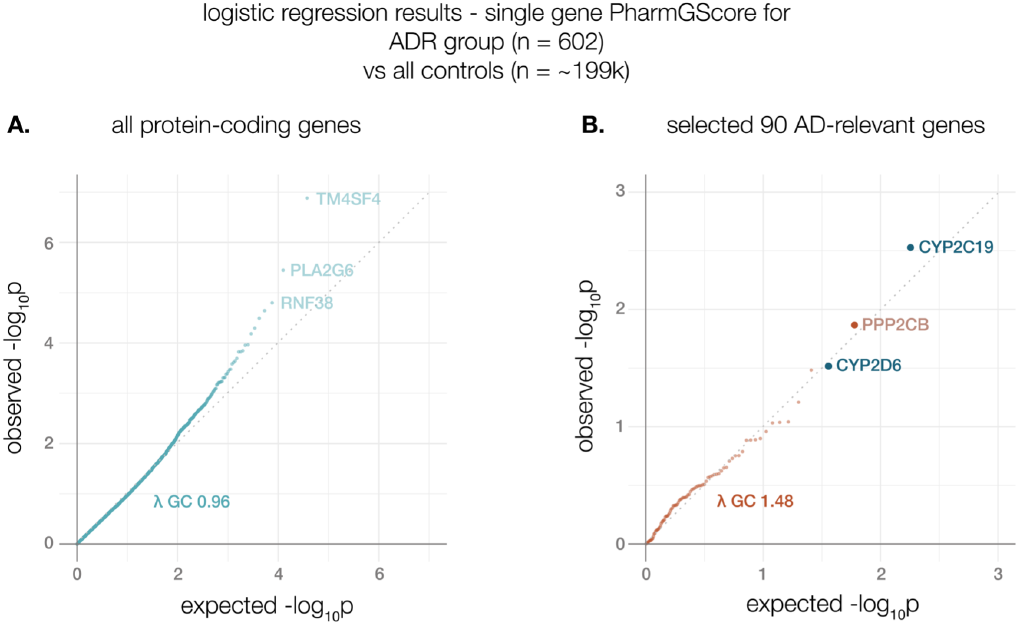
Quantile-quantile (qq) plots of p-value distributions from logistic regression comparing the ADR group with all controls. A. qq plot for logistic regression results for the ADR phenotype on all protein-coding genes, three top genes are labeled (these passed the FDR threshold of 10%). B. Qq plot for the 90 selected genes, including 9 cytochrome P450 genes. The top 3 genes are labeled. *CYP2C19* and *CYP2D6* have a distinct color as these are cytochrome P450 genes already indicated in antidepressant responses. Additional plots for other comparisons are available in the supplementary material (supplementary figure 4). Full statistical results are available in supplementary table G.

### Compound PharmGScore prediction of ADRs

To compare compound PharmGScore (Figure 4A) one-way ANOVA with Tukey’s post-hoc test was used. To analyze the compound PharmGScore association with accidental poisoning/adr (described in the last paragraph of the results section) Walds’ logistic regression test was used. To assess the classification ability of the compound PharmGScore of accidental poisonings based on different gene lists, we plotted its ROC curves (Figure 4B, Supplementary Figure 5) and compared each with 1000 ROCs based on randomly drawn gene sets with the same number of genes. The AUC p-values were estimated with empirical distribution function.

**Figure 4.**
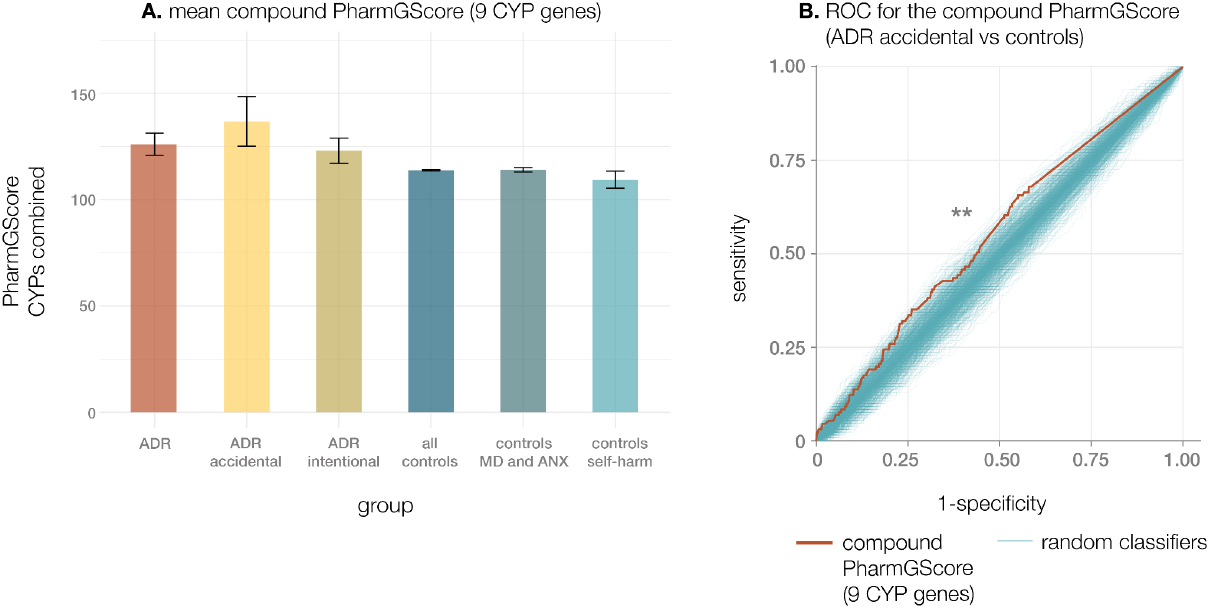
Performance of the compound PharmGScore from 9 cytochrome P450 (CYP) genes (*CYP2C19, CYP2C9, CYP2D6, CYP3A5, CYP2B6, CYP1A2, CYP2C8, CYP3A4, CYP2E1*) in participants with antidepressant toxicity/ADR diagnosis. The compound PharmGScore is defined as the sum of scores for all 9 genes. Only genes with PharmGScore > 50 are summed. A. Comparison of the mean PharmGScore across groups. Statistical differences were analyzed with one-way ANOVA with Tukey’s post-hoc test; no comparisons passed the corrected p-value threshold 0.05 - results are available in the supplementary table H). B. ROC curve of a classifier distinguishing participants of the “ADR accidental” group (n = 131) versus all controls (n = 199691) showing classification performance of the compound PharmGScore based on 9 cytochrome P450 genes (red thick line). For comparison, 1000 ROCs of sets of 9 randomly drawn protein-coding genes are shown (blue thin lines). The AUC of the classifier based on 9 CYP genes is 0.55 and empirical p-value of this model is = 0.009.

## Results

### PharmGScore burden score validation

Our goal was to build an ensemble score with a focus on pharmacogenes and their functional impact that would allow for combining multiple variants and genes into one value. We validated the score using pharmacogenetic variant definitions deposited in PharmVar. For each variant definition we calculated the PharmGScore for its component SNVs and summed it. The average number of SNVs included in each star allele was 5.16. Overall 374 star allele definitions were tested from 8 genes and included 326 unique single nucleotide variants (Supplementary Table B).

We compared the PharmGScore with the scores it was built from and show its increased performance for pharmacogenetic variants (Figure 1A). As the main assessment we evaluated the ability of the score to differentiate between normal and increased function variants versus decreased and no function variants (ROC AUC: PharmGScore = 0.86, CADD = 0.79, FATHMM-XF = 0.81, PROVEAN = 0.81, Mutation Assessor = 0.75). This evaluation considered variants within 50 bp from each exon. Inclusion of less (distance to exon = 0 bp) or more (distance to exon = 100 bp) non-coding variants did not improve the performance of the star allele PharmGScore (Supplementary Figure 1). We additionally investigated the PharmGScore ability to differentiate between all star allele subtypes (Supplementary Figure 2) and found that the score performs the best in the classification of no function variants (AUCs: no function vs normal function = 0.86, no function vs increased function = 0.89), well with the decreased function (AUCs: decreased function vs normal function= 0.86; no function vs decreased function = 0.70; increased function vs decreased function = 0.96) and has the lowest performance in differentiating increased function variants from normal function (AUC normal function vs increased function=0.56.

We compared PharmGScore values for five antidepressant-relevant cytochrome P450 (CYP2) genes individually and found that the no function star alleles have a significantly higher PharmGScore in four out of five genes after correction for multiple comparisons (Figure 1B). Decreased function star alleles have lower median burden scores than no function alleles and for *CYP2B6* and *CYP2D6* this difference was statistically significant (Figure 1B). To determine the PharmGScore cut-off point we aimed for a threshold with a low false positive rate. The rationale behind this was to prevent the false positives from overwhelming the compound PharmGScore (summed from multiple genes). We, therefore, determined the PharmGScore cut-off point to be 50 for each star allele and gene. In that manner, the false positive rate of PharmGScore is equal to 0.6% based on all PharmVar star allele definitions. At this cut-off 20.8% of decreased function and 59.2% of no function PharmVar star alleles for cytochrome P450 genes (CYP) are above the threshold (Figure 1B).

### Annotation of the UKB Whole Exome Sequencing data with the PharmGScore

To explore the association between PharmGScore and real-life phenotypes we annotated the UK Biobank’s 200k Whole Exome Sequences (WES) with the summed genel-level PharmGScore for each of the protein-coding genes. We first focused our analyses on cytochrome P450 (CYP) genes. The gene-level PharmGScore distribution for two of the cytochrome P450 genes from the whole 200k WES cohort is shown in Figure 2A and B. As described above, the cut-off threshold of 50 was used to determine the gene PharmGScore to be high. To investigate the burden of all selected 9 cytochrome P540 (CYP) genes we additionally calculated a compound PharmGScore, where scores for genes with PharmGScore above 50 were summed for all nine genes and the threshold was set at 100. This value was chosen for the score to be above the threshold if a participant had at least two genes with PharmGScore > 50. For this score, around 40% of participants were above the threshold (Figure 2C).

### A higher PharmGScore in cytochrome P450 genes is associated with a higher odds ratio of antidepressant toxicity

After obtaining PharmGScores for each of the genes for the UKB participants we aimed to assess if a high burden in selected genes is associated with psychiatric drug toxicity codes. We included all relevant codes that had more than 200 sequenced cases: poisoning by other and unspecified antidepressants (T43.2), poisoning by benzodiazepines (T42.4), poisoning by other opioids (T40.2), opioids and related analgesics causing adverse effects in therapeutic use (Y45.0) and poisoning by other antiepileptic and sedative-hypnotic drugs (T42.6).

We analyzed high gene-level (>50) and compound (>100) PharmGScore of cytochrome P450 genes in association with the occurrence of toxicity and ADR codes related to the central nervous system. We found that a higher burden in *CYP2C19, CYP2C9* and a compound score from all nine cytochrome P450 genes was associated with a higher Odds Ratio of poisoning by unspecified antidepressants (Table 1, Supplementary table E). To investigate if the results can be generalized to all genetic backgrounds we also performed the chi2 test for the compound PharmGScore from 9 cytochrome P450 genes in non-European participants. It did not achieve statistical significance, but the direction of changes was the same as in European UKB participants (p-value 0.21; odds ratio 1.44, 95% CI for odds ratio: 0.81-2.56). Results for other diagnostic codes studied did not show any significant overrepresentations (Supplementary table E). Apart from cytochrome P450 (CYP) genes, membrane transporters, receptors and other genes may also play a role in antidepressant ADR. Therefore we performed the same tests for 81 additional relevant genes, but found no significant associations (Supplementary Table E).

### Antidepressant toxicity is more common in participants with high PharmGScore in selected genes

Above-described analysis showed that higher PharmGScore in pharmacogenes was associated with a higher odds ratio of an antidepressant toxicity code. To get more insight into the pharmacogenetics of antidepressant drugs we studied the per-gene PharmGScore in the group of UKB participants that received any antidepressant toxicity or adverse drug reaction diagnosis in the hospital codes. This group, shortly termed here as the ADR group, consisted of 602 individuals. Notably, the majority of the poisonings were intentional (see Supplementary Table F for detailed characteristics). To control for the effect of self-harm tendencies and mental health conditions we compared the ADR group with additional control groups: participants with a history of self-harm with drugs and participants with hospital codes of depression and/or anxiety. ADR group subjects had a higher Townsend deprivation index, proportionally more females and were younger than the average UKB participant (Supplementary Table F). Age, sex and Townsend index were included as covariates in the variant burden analyses (together with the first 17 genetic PCs).

To evaluate the relationship between PharmGScore variant burden in individual genes and ADRs to antidepressants or antidepressant poisoning we performed logistic regression analyses on 1) all protein-coding genes (n = 19342) and 2) the antidepressant-response relevant genes (9 cytochrome P450 and 81 additional genes, gene list available in Supplementary Table D). We ensured that our model did not inflate statistical significance by drawing a graph of expected vs observed p-values (qq plot). The genomic inflation factor lambda (Figure 3A) was below the assumed threshold of 1.05 therefore we concluded that the p-values were not inflated. On the whole genome scale, 3 genes passed the 10% FDR threshold: *TMS4SF4, PLA2G6, RNF38*. We then compared the results for these genes with logistic regression results from other comparisons (Table 2). High PharmGScore in *PLA2G6* remained associated with antidepressant toxicity/ADR if controls were restricted only to subjects suffering from depression and/or anxiety. There were no differences in the PharmGScore for *TMS4SF4, PLA2G6, RNF38* in the accidental vs intentional poisoning comparison. In comparison with the group that had self-harm history with drugs none of the three genes remained significant (the FDRs for *TMS4SF4, PLA2G6, RNF38* were close to 1 with *PLA2G6* having the lowest p-value).

Our main aim in this part of the study was to interrogate variant burden in genes known or suspected to contribute to antidepressant drug responses (n = 90 genes), including 9 cytochrome P540 (CYP) genes). For this geneset, we observed an overall inflation of p-values from the expected distribution with the inflation factor 1.48 (Figure 3B). No genes passed the assumed 10% FDR threshold in this analysis. However, known antidepressant-relevant pharmacogenes: *CYP2C19* and *CYP2D6* were in the top three genes, together with the *PPP2CB* gene discovered in an antidepressant response sequencing study (Xu et al. 2020). Both *CYP2D6* and *CYP2C19* were in the top three genes with the lowest p-values for comparisons between the ADR group with all controls, including controls with self-harm history (Table 3). Furthermore, they were not in the top 3 genes (32nd and 48th respectively) for the comparison between accidental and intentional poisonings.

### Predicting ADRs to antidepressants with compound PharmGScore

To assess the possibility to predict any type of Adverse Drug Reaction or antidepressant toxicity with computational burden scores we performed analyses of the compound burden from all 9 cytochrome P450 genes for the ADR group. To test the robustness of the metric we used a single compound PharmGScore for each individual, without any exclusions or adjustment. First, we compared the mean compound PharmGScore for all the nine cytochrome P450 genes between the ADR group, the accidental and intentional poisoning subgroups and all the control groups (Figure 4). The mean compound PharmGScore was lower for all the control groups than all the ADR groups and was, on average, the highest in the group with accidental poisonings, however, none of the t-test p-values passed the 0.05 threshold after Bonferroni correction (Supplementary table H).

We then conducted additional tests to investigate the compound PharmGScore in the group of participants with accidental poisonings / ADRs with therapeutic doses (n = 131). Firstly, we performed a logistic regression test for the ADR-accidental phenotype for the whole cohort, without considering covariates. We found that higher compound PharmGScore is indeed associated with a higher likelihood of unintentional antidepressant toxicity/ADR (beta = 0.0014, p-value 0.035; full results available in Supplementary Table I). The test was then repeated for non-European participants and the association had the same direction but was no longer significant (beta = 0.00169, p-value 0.49; full results available in Supplementary Table I).

Finally, we assessed the performance of classifiers based on compound PharmGScore in distinguishing controls from participants with unintentional antidepressant poisonings and ADRs at therapeutic doses. We found that a classifier based on selected 9 CYP genes performed statistically significantly better than a set of control models at distinguishing patients from controls (Figure 4B, p-value = 0.009, AUC = 0.55). To investigate the value of adding additional relevant genes we evaluated a classifier based on all 90 selected genes, which obtained a p-value of 0.12 and performed better than any other classifier at 80% specificity (Supplementary Figure 5).

## Discussion

Due to the lack of novel pharmacotherapies there is an urgent need for psychiatric treatment personalization and pharmacogenetics offers a way to optimize the acceptability of psychiatric drugs. This study aimed to show that exome sequencing data and computational variant burden prediction could become valid methods for pharmacogenetic assessment. Our main finding is that a computational variant burden score can be applied to real-life healthcare data and demonstrate the known contribution of cytochrome P450 genes to antidepressant toxicity. The main novelty of this approach is the ability to depart from the star allele nomenclature and apply a single score for pharmacogenetic prediction.

### The PharmGScore strengths, limitations and prospects

Traditionally the pharmacogenetic haplotypes were assigned to star alleles, and experiments were performed to assess the enzyme function. However, with genome sequencing data it has become unfeasible to investigate each allele separately as the genotype discovery outpaces the capability of functional studies (Brown, Staples, and Woodahl 2022). Machine learning techniques that offer prospects for the prediction of protein function are in development (McInnes et al. 2020; Pandi et al. 2021). Another approach is to leverage computational prediction of genetic variant impact via available tools. It has been previously shown that scores optimized for the whole genome do not have sufficient sensitivity and specificity (Zhou et al. 2019). Current genetic variant scores have several limitations in regards to pharmacogenetics: they have not been tested on known pharmacogenetic variants and are difficult to compare and combine between genes. Previously, using an average of multiple variant scores proved to be useful to assess the impact of single pharmacogenetic variants (Zhou et al. 2019).

The developed PharmGScore includes four component scores, which are all established and well-validated. It involves per-gene normalization, which takes into account varying gene lengths, and has higher predictive performance than its component scores. PharmGScore, due to the normalization step, allows combining multiple genes into a single score. Here we combined the PharmGScore for nine cytochrome P450 genes indicated in antidepressant drug responses. The metric is currently based on WES data, which means that not all non-coding mutations are included. However, the majority of variants recorded in PharmVar are actually captured by WES. In the current version of the PharmGScore we included a region of 50 bp before and after each exon. This way 85% of all SNVs from PharmVar are included (90% for genes other than *CYP2D6*). Moreover, our data indicate that inclusion of variants further than 50 bp from each exon does not improve the score’s performance. Careful incorporation of whole genome sequences as they become more available is one of the future directions of PharmGScore development.

The PharmGScore is publicly available and can, in principle, be used to assess genetic mutational burden in any genes and gene sets for any pharmacological traits. Another advantage is that it incorporates all genetic variants of a patient no matter the population frequency and therefore leverages sequencing data to the full extent. This is relevant, as rare genetic variants contribute substantially to the functional variation of cytochrome P450 (CYP) genes(Zhou et al. 2022). Limitations of the score include its relatively low sensitivity for decreased protein function at the assumed threshold of 50. However, the overall performance of the PharmGScore was 86% for differentiating between normal and decreased function star alleles and 70% for distinguishing between no and decrease function variants. Moreover, the score is currently detecting predominantly deleterious mutations and is not fine-tuned to detect increased function variants. With further refinement, the score could better resolve distinct consequences of mutations, such as increased or decreased function.

### The application of the PharmGScore to the UK Biobank data

The prospects of the PharmGScore for treatment optimisation were investigated in participants of the UKB that received antidepressant toxicity or ADR diagnoses. Whenever possible, we included the whole 200k WES cohort, without excluding any ethnicity. This is important as non-European populations carry a greater proportion of deleterious pharmacogenetic variants and thus may be more prone to drug toxicity (McInnes et al. 2021). We performed statistical tests in the non-European participants’ subgroup and, although the test results were not significant, found the same direction of changes as for the whole cohort. Notably, the phenotypic data collected by the UKB is inherently noisy as it was collected in a real-life healthcare setting. Using such datasets to investigate pharmacogenetics gives insight into how developed tools may perform in the clinic.

### The pharmacogenetics of AD toxicity and ADRs: role of cytochrome P450 genes and other genes

Genotypes in *CYP2D6* and *CYP2C19* are known to be associated with a higher risk of ADRs, although not much is known about the genetics of overdoses and serious ADRs to antidepressants. ADRs to antidepressant drugs cause around 25k Emergency Room visits each year in the United States alone and include seizures, toxic delirium, collapses, neurological ADRs, hyponatremia and allergic reactions (Degner et al. 2004; Hampton et al. 2014). A recent study of the association of *CYP2C19* metabolizer status in a clinical setting and ADRs to antidepressants in therapeutic doses found that Poor Metabolizers (PMs) had a higher risk of autonomic and neurological side effects, although the improvement of their symptoms was also higher (Calabrò et al. 2022). A meta-analysis of GWAS confirmed that *CYP2C19* PMs had higher odds of antidepressant ADRs in the first 2 to 4 weeks of treatment (Chiara Fabbri et al. 2018). *CYP2D6* PMs also tend to have higher chances of ADRs to amitriptyline, nortriptyline, venlafaxine, and fiuoxetine (Haufroid and Hantson 2015).

In our study, there are two major considerations to take into account when interpreting the results. Firstly, due to the limited information available, we did not consider antidepressants or antidepressant drug classes separately, which would likely have brought more precision to the results. This is a common approach in genetic studies due to limited group sizes and significant associations with antidepressant responses and genetic factors have been previously discovered in this manner (Cocchi et al. 2016; Xu et al. 2020). Secondly, in the cohort of patients that have been assigned ADRs to antidepressants as hospital codes, the vast majority are serious reactions due to intentional overdoses. There are indications that severe consequences (e.g. hospitalization, death) of intentional overdoses may be more common in carriers of particular pharmacogenetic variants. Individual cases of defective *CYP2D6* genotypes in fatal amitriptyline and doxepin poisonings are described in the literature (Koski et al. 2006, 2007). A study of 349 suicides of citalopram users found that combined together, poor and ultrarapid metabolisers of *CYP2C19* were overrepresented in the suicide group (Rahikainen et al. 2019). In this study, we mitigated the issue of intentionality in two ways. Firstly, in the logistic regression analysis, we included an additional control group that consisted of UKB participants with a history of self-harm with drugs. Known antidepressant-relevant genes (*CYP2D6* and *CYP2C19*) remained within the top genes when the ADR group was compared with this control group, while p-values for other genes (e.g. *TM4SF4, RNF38*) dropped considerably. Secondly, we confirmed that the compound PharmGScore predicts accidental poisonings or severe ADRs that occurred at therapeutic doses.

Here we demonstrate that participants with a higher gene-level PharmGScore in *CYP2C19, CYP2C9* are overrepresented with the patient group assigned the toxicity of other and unspecified antidepressants at an FDR threshold of 0.1. *CYP2C19* was also among the top 3 most significant genes (when a preselected list of 90 genes was considered) for the comparisons between all participants in the ADR group and all selected controls. In the intentional vs accidental poisonings comparison, the *CYP2C19* gene ranked in the 2nd half of all p-values. This indicates that the deleterious variants in *CYP2C19* may contribute to hospital visits due to antidepressant toxicity regardless of the intention. It is worth noting that, likely due to insufficient statistical power, the logistic regression results for *CYP2C19* do not meet the corrected p-value significance threshold. Therefore the relationship between *CYP2C19* deleterious variants and antidepressant responses should be further researched in the context of intentional and accidental poisonings.

*CYP2D6* is known to carry multiple structural variants, which were not taken into account in this study (Nofziger et al. 2020). A further refinement of the PharmGScore that would include *CYP2D6* structural variant analysis could increase the predictive power of the algorithm. However, in clinical settings, often no structural variant data is available. Therefore is it valuable to assess the score performance on *CYP2D6* with only single-nucleotide variants included. Here we show that in a list of 90 genes *CYP2D6* is within the top 3 genes in the comparison of the ADR group with all controls even if only single nucleotide variants are considered. Importantly, *CYP2D6* was the top gene in the comparison of the ADR group with the control group with self-harm with drugs history, which indicates that, as with *CYP2C19*, deleterious variants in *CYP2D6* contribute to antidepressant toxicity also when the drug is intentionally overdosed.

*CYP2D6, CYP2C9* and *CYP2C19* are not the only cytochrome P450 genes that metabolize antidepressants. PharmGKB lists also *CYP3A5, CYP2B6, CYP1A2, CYP2C8, CYP3A4* and *CYP2E1* in relation to distinct antidepressant classes (Whirl-Carrillo et al. 2021). In our analyses we assessed not only single-gene PharmGScore but also a compound score built from 9 genes. A larger compound PharmGScore was associated with a higher probability of being in the ADR group. This association held both if demographic, genetic and socioeconomic covariates were included in the test and when they weren’t. Moreover, although the statistical results for non-European participants were not significant, the direction of observed associations was the same as in the European group. These findings indicate that PharmGScore may generalize to distinct patient groups and populations, which is of importance to the clinic. Although in our tests we did not observe any differences between intentional and accidental poisonings, the accidental ADR group had, on average, the highest compound PharmGScore suggesting that cytochrome P450 deleterious genotypes have the highest contribution to hospitalization in this group.

Apart from cytochrome P450 genes we investigated other genes that may play a role in antidepressant responses. In the exploratory analysis of all protein-coding genes three passed the 10% FDR threshold: *TM4SF4, RNF38* and *PLA2G6. TM4SF4* and *RNF38* ranked very low (1216 and 1424 respectively) in the test comparing the ADR group with participants that had self-harm with drugs in history. This indicates that deleterious variants in these genes may be associated with self-harm tendencies. Mutations in the epithelial *TM4SF4* are typically associated with gallstones (Weber et al. 2019) and have not been reported in mental disorders thus far. However, the expression of *TM4SF1*, a paralog of *TM4SF4*, was found to be lower in completed suicide cases with bipolar disorder (Kim et al. 2007). Variants in *RNF38* are associated with autism (Abrahams et al. 2013), but were not reported to date with regard to antidepressants, depression or self-harm tendencies. *PLA2G6* gene was significant (FDR < 0.001) in the comparison of the ADR group with controls suffering from depression and/or anxiety and ranked 36th in the comparison of the ADR group along the self-harm axis. *PLA2G6* encodes Phospholipase A2 and is associated with neurodegeneration phenotypes (Gregory et al. 2017) and should be considered as a potential candidate gene associated with ADRs to antidepressants.

In the analysis of the preselected gene set (90 genes including 9 CYP genes) we observe inflation of the p-values which indicates that this list is enriched in genes with higher PharmGScore associated with ADRs to antidepressants. Apart from the cytochrome P450 genes, *PPP2CB* was in the top three genes when all controls were compared to the ADR group (although it did not pass FDR). It was the 3rd and 18th gene when ADR group was compared with controls with depression and/or anxiety and self-harm tendencies respectively. The *PPP2CB* gene, encodes a subunit of the serine/threonine-protein phosphatase 2A, was identified as a gene associated with antidepressant responses in an exome sequencing study (Xu et al. 2020) and here we provide further indications that it may play a role in antidepressant toxicity.

One of the aims of this study was to expand the list of genes included in pharmacogenetic analyses. The performance of the compound PharmGScore based on 9 cytochrome P450 genes indicates that antidepressant pharmacogenomics benefit from the addition of genes outside of *CYP2D6* and *CYP2C19*. We further investigated a list of additional genes from the literature and we did not observe significant associations of gene PharmGScore with antidepressant toxicity. However, a classifier based on a list of 90 genes performed both better than any of the control classifiers and better than the CYP-based classifier at high specificity thresholds (e.g. 80%). This is consistent with the idea that damaging variants in genes other than cytochrome P450 influence antidepressant responses. However, when all protein-coding genes were included the classifier performed poorly, with an overall AUC below the chance level (AUC = 0.49). We therefore conclude that inclusion of relevant genes from the literature, with further testing, may be beneficial for pharmacogenetic testing. However, based on the current state of knowledge, including all available genetic information may be suboptimal for drug response prediction.

## Conclusions

With this study we show, for the first time, how a single genetic burden score may be associated with the toxicity of psychiatric drugs recorded in healthcare data. The analyses demonstrated that even intentional toxicity cases reported in hospitals tend to have a higher genetic burden in known pharmacogenes therefore it is valuable to include such cohorts in pharmacogenetic studies. Furthermore, our results confirmed that *CYP2C19* and *CYP2D6* are among the top genes associated with antidepressant drug responses without using the traditional star allele calling but rather a computational predictor. We present the first iteration of a framework that, with refinement and development, could serve as an approach complementary to star allele genotyping. Even at the current stage, the PharmGScore can be used as an additional tool to indicate patients that may be more susceptible to overall antidepressant toxicity. The metric is freely available and can be used in any pharmacogenetic research where exome or genome sequences are available. There are no restrictions on the drug types the PharmGScore can be applied to, although separate validation for each drug is necessary.

## Data Availability

All the code used in this project is available in a GitHub repository (https://github.com/ippas/ifpan-pharmgscore-manuscript). The star allele definitions were downloaded from the Pharmacogene Variation Consortium (PharmVar v 5.1.6) at www.pharmvar.org (Gaedigk et al. 2021). The component scores of the PharmGScore are publicly available: CADD (Rentzsch et al. 2019), FathmmXF (Rogers et al. 2018), PROVEAN (Choi and Chan 2015) and MutationAssessor (Reva, Antipin, and Sander 2011).
The UK Biobank data used in this research is available to approved researchers and has been accessed through the application 62979 Impact of pharmacogenetic profiles on depression treatment outcomes. The pre-computed PharmGScore for all possible single-nucleotide variants (SNV) in WES is available for download as tsv: https://pharmgscore.labpgx.com/ (Supplementary Table A)

https://pharmgscore.labpgx.com/

https://github.com/ippas/ifpan-pharmgscore-manuscript

## Acknowledgements

This research has been conducted using the UK Biobank Resource under Application Number 62979. We are grateful to the UK Biobank and all its voluntary participants. This work used data provided by patients and collected by the NHS as part of their care and support. The UK Biobank study was conducted under generic approval from the NHS National Research Ethics Service (approval letter dated 17 June 2011, Ref 11/NW/0382). Copyright © 2023, NHS England. Re-used with the permission of the UK Biobank. All rights reserved. This research also used data assets made available by National Safe Haven as part of the Data and Connectivity National Core Study, led by Health Data Research UK in partnership with the Office for National Statistics and funded by UK Research and Innovation (grant: MC_PC_20029). We would also like to thank Dr Chiara Fabbri for her valuable comments on the manuscript.

## Funding

The research in this manuscript was funded by the National Science Center grant SONATINA UMO-2021/40/C/NZ2/00218 awarded to MB and the statutory funds of the Maj Institute of Pharmacology Polish Academy of Sciences. The computational part of this research was supported by PLGrid Infrastructure.

## Authors contribution

MK, MP and MB designed the research, JH conducted the analyses, MB and JH prepared the figures and tables, MB drafted the manuscript, JB wrote parts of the manuscript, all coauthors revised the manuscript and provided essential input into the final version.

## Conflict of Interest statement

The authors declare that the research was conducted in the absence of any commercial or financial relationships that could be construed as a potential conflict of interest.

## Notes

### Competing Interest Statement

The authors have declared no competing interest.

### Author Declarations

The UK Biobank study was conducted under generic approval from the NHS National Research Ethics Service (approval letter dated 17 June 2011, Ref 11/NW/0382).

