## supplemental material for "PharmGScore scores of compound genetic variant burden for psychiatric treatment optimization"

**Supplementary table A**. PharmGScore for every single nucleotide variant within 50bp range from exons of a gene. Available to download in the .tsv format: https://pharmgscore.labpgx.com/

**Supplementary table B**. Star alleles and variants included in them used in the PharmGScore validation process

**Supplementary table C**. Statistical results for figure 1.

**Supplementary table D**. Genes selected as relevant for AD responses analyzed in this study. First 9 genes are cytochrome P450 genes (column A), and the rest are diverse genes used as the extended gene set (column B).

**Supplementary table E**. Full results of diagnostic code overrepresentation analysis for 9 CYP genes and their composition (sheet A) and the extended gene set (sheet B).

**Supplementary table F**. Phenotypic characteristics of the ADR group and controls.

**Supplementary table G.** Full results of all the logistic regression analyses comparing the ADR group to various controls. Results are divided into two sections: the selected genes and all protein coding genes. The FDR was applied within the geneset, each sheet is named with the comparison and geneset it includes. **AD-relevant** - 90 genes preselected from the literature; **all** - all protein coding genes; **adr-control** - comparison of the ADR group with all controls; **intention** - the intentional vs accidental poisonings; **mdanx** - the ADR group compared with other UKB participants that received anxiety and or depression diagnosis; **self-harm** - the ADR group compared with UKB participants that had a history of self-harm with drugs.

**Supplementary table H**. Statistical results for figure 4.

**Supplementary table I**. Logistic regression results for the ADR-accdidental phenotype based on PharmGScore composed from 9 cytochrome P450 genes.

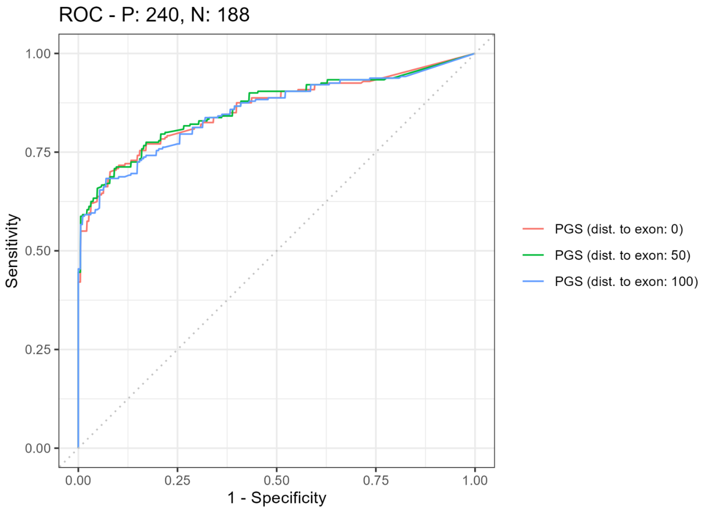


**Supplementary Figure 1.** ROC curves for PharmGScore depending on the distance to exon in which variants were included (0, 50 or 100 bp). For this ROC normal and increased function star alleles were assigned a value of 0, while no function and decreased function alleles were assigned a value of 1. AUCs of the PharmGScore: dist. to exon: 0 = 0.859, dist. to exon: 50 = 0.863, dist. to exon: 100 = 0.854

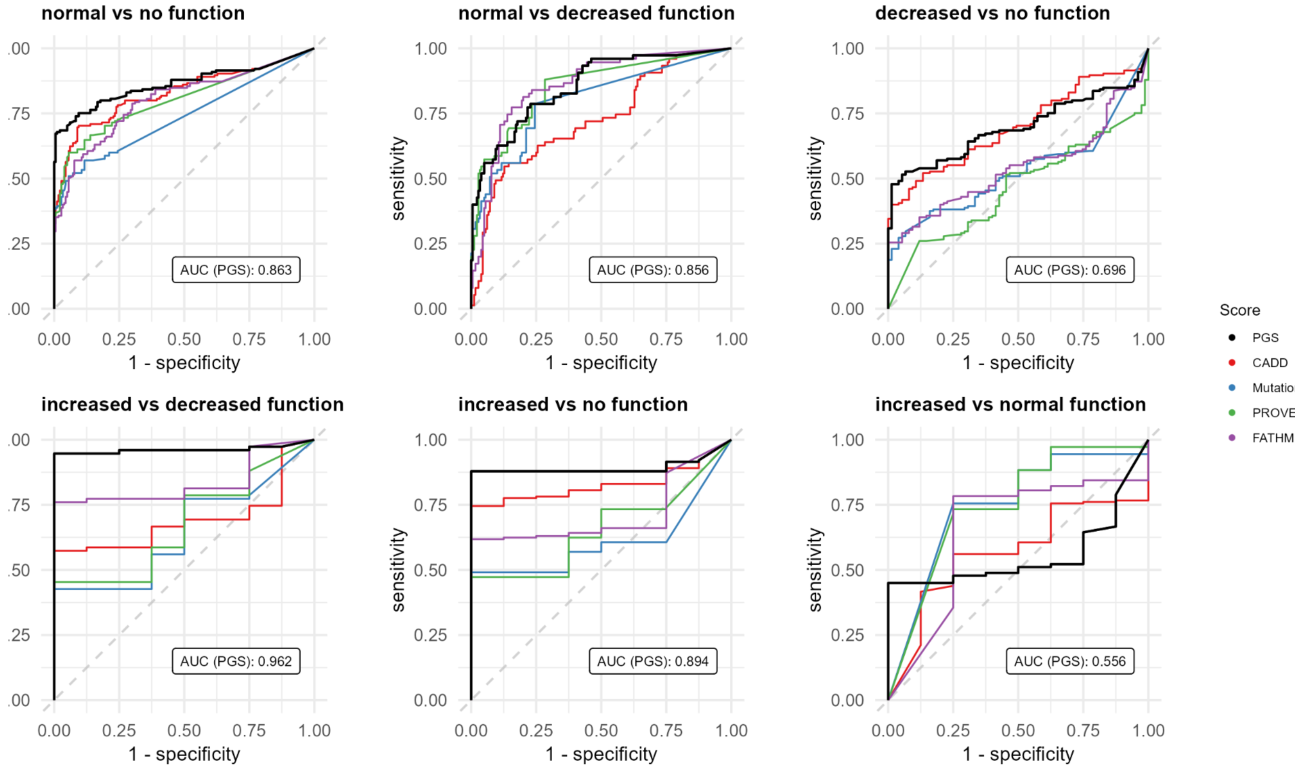


**Supplementary figure 2.** ROC curves for the PharmGScore and its component scores in classification performance between each star allele category. AUCs for each function pair for PGS score are displayed within the plot. There are only 8 increased function variants in the PharmVar database, therefore the curves on the three lower plots have a jagged appearance. AUCs for each function pair and each compound score: normal vs no function: PharmGScore = 0.863, CADD = 0.828, Mutation Assessor = 0.737, PROVEAN

= 0.801, FATHMM-XF = 0.801; normal vs decreased function: PharmGScore = 0.856, CADD = 0.726, Mutation Assessor = 0.802, PROVEAN = 0.851, FATHMM-XF = 0.858; decreased vs no function: PharmGSCore = 0.696, CADD = 0.699, Mutation Assessor = 0.531, PROVEAN = 0.463, FATHMM-XF = 0.543; increased vs decreased function: PharmGScore = 0.962, CADD = 0.692, Mutation Assessor = 0.647, PROVEAN = 0.675, FATHMM-XF = 0.835; increased vs no function: PharmGScore = 0.894, CADD = 0.828, Mutation Assessor = 0.608, PROVEAN = 0.656, FATHMM-XF = 0.714; increased vs normal function: PharmGScore = 0.556, CADD = 0.568, Mutation Assessor = 0.748, PROVEAN = 0.748, FATHMM-XF = 0.655.

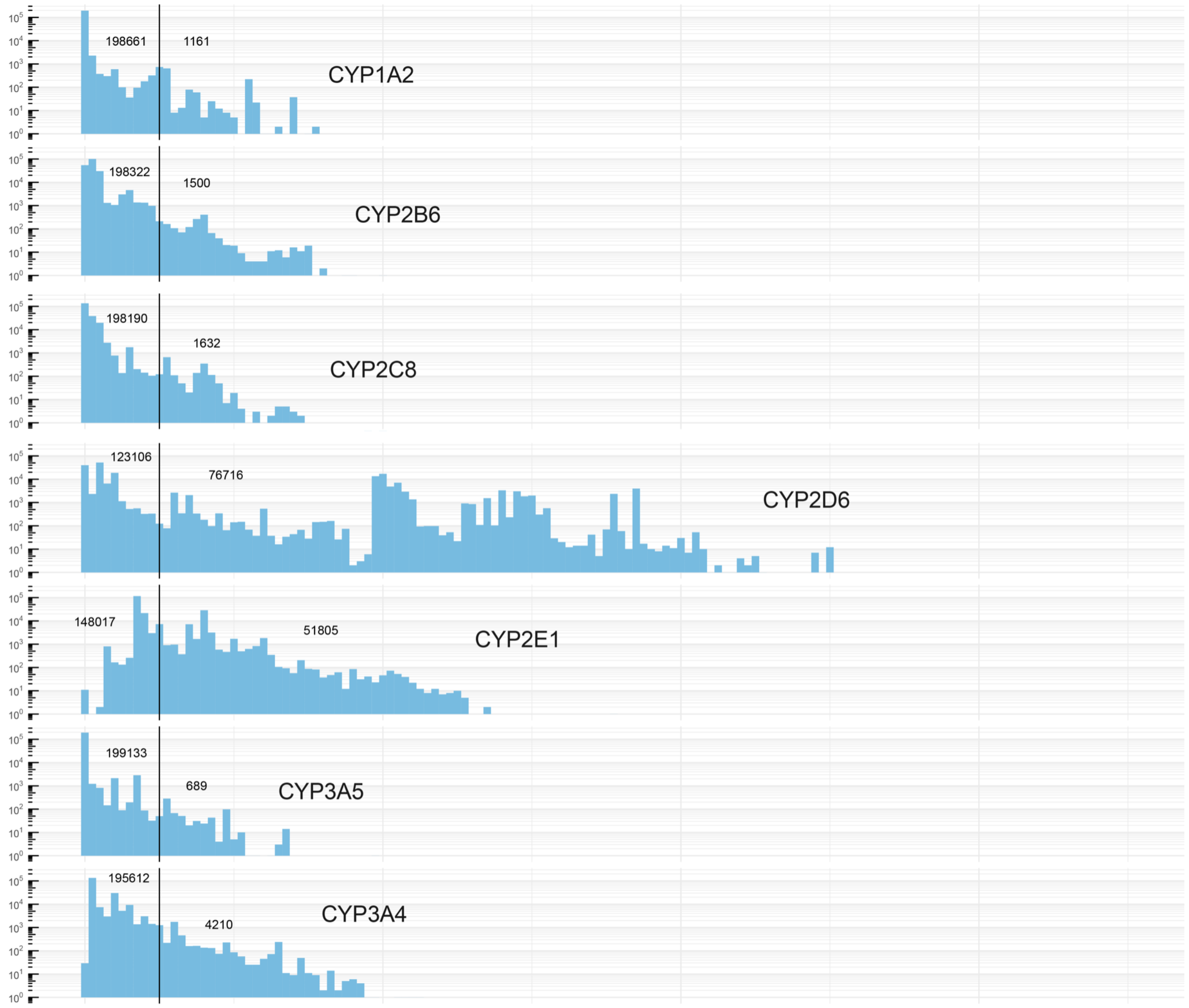


**Supplementary Figure 3.** Distribution of the PharmGScore in the 200 UKB WES for the remaining CYP genes not shown in Figure 2. X axis - summed PharmGScore value; Y axis - number of participants (log10-scaled).

**A.**

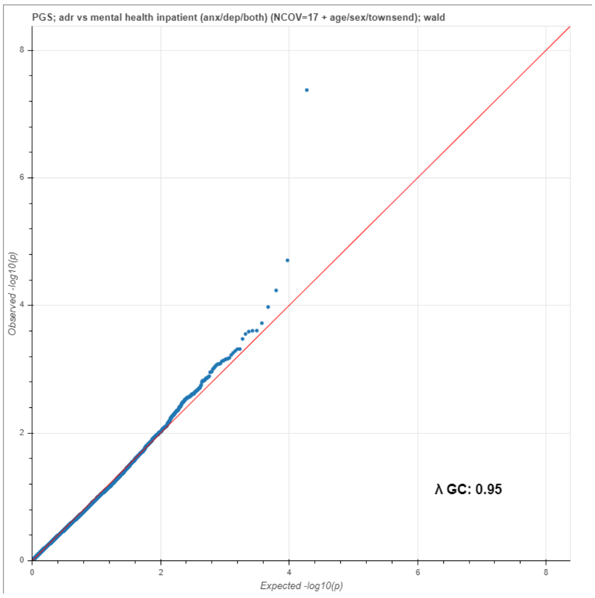
PGS; adr vs mental health inpatient (anx/dep/both) (NCOV=17 + age/sex/townsend); wald
Observed -log10(p)
Expected -log10(p)
λ GC: 0.95

**B.**

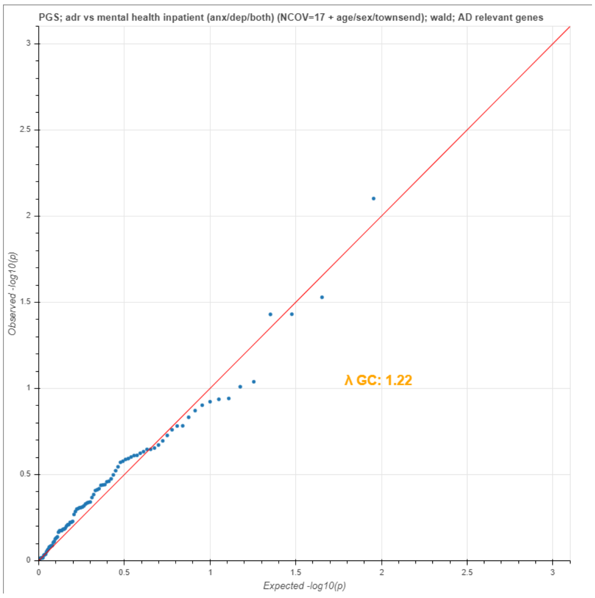
PGS; adr vs mental health inpatient (anx/dep/both) (NCOV=17 + age/sex/townsend); wald; AD relevant genes
Observed -log10(p)
Expected -log10(p)
λ GC: 1.22

**C.**

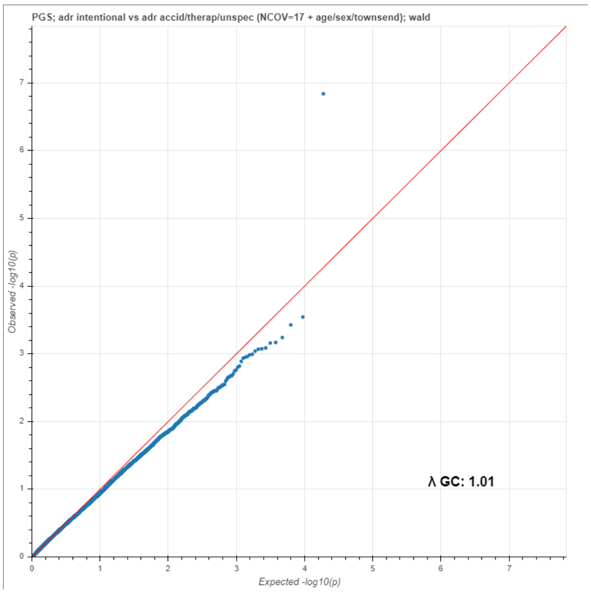
PGS; adr intentional vs adr accid/therap/unspec (NCOV=17 + age/sex/townsend); wald
Observed -log10(p)
Expected -log10(p)
λ GC: 1.01

**D.**

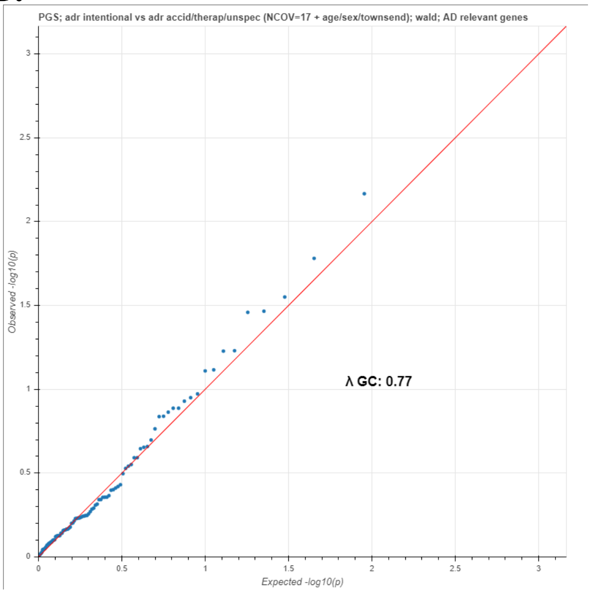
PGS; adr intentional vs adr accid/therap/unspec (NCOV=17 + age/sex/townsend); wald; AD relevant genes
Observed -log10(p)
Expected -log10(p)
λ GC: 0.77

**E.**

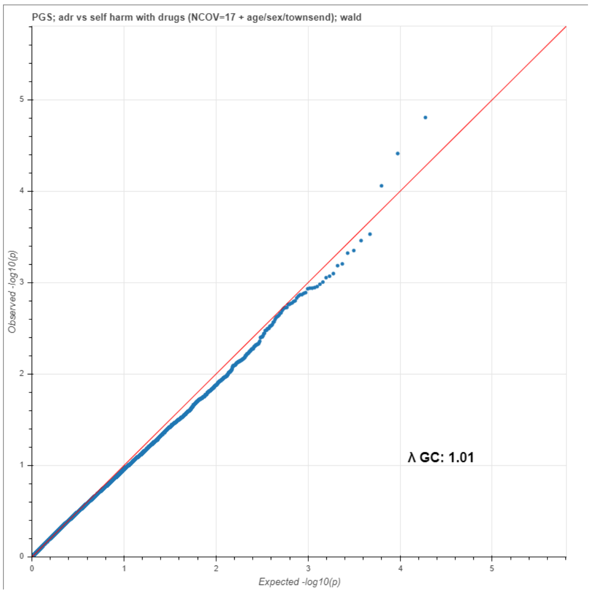
PGS; adr vs self harm with drugs (NCOV=17 + age/sex/townsend); wald
Observed -log10(p)
Expected -log10(p)
λ GC: 1.01

**F.**

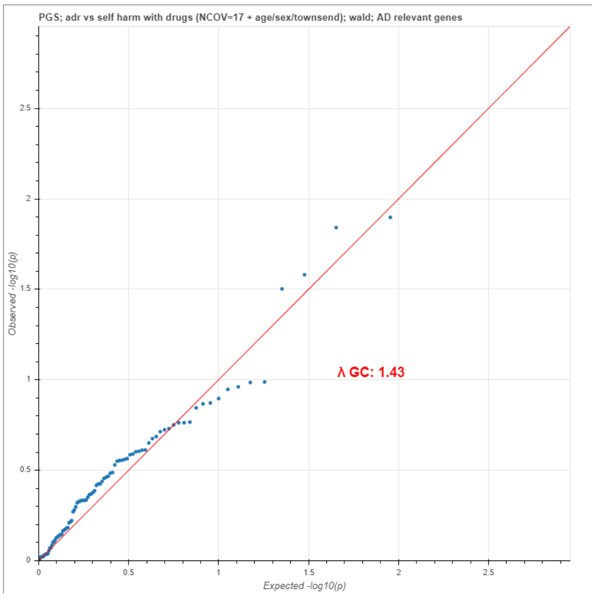
PGS; adr vs self harm with drugs (NCOV=17 + age/sex/townsend); wald; AD relevant genes
Observed -log10(p)
Expected -log10(p)
λ GC: 1.43

**Supplementary figure 4.** Qq plots for all protein coding genes and the 90 AD relevant genes for linear regression comparing the ADR group to various control groups. A. All genes for ADR vs participants with depression and/or anxiety, B. AD-relevant genes ADR vs participants with depression and/or anxiety, C. All genes comparison between accidental and intentional poisonings, D. AD-relevant genes comparison between accidental and intentional poisonings , E. All genes comparison between ADR and participants with self-harm with drugs history, F. AD-relevant comparison between ADR and participants with self-harm with drugs history

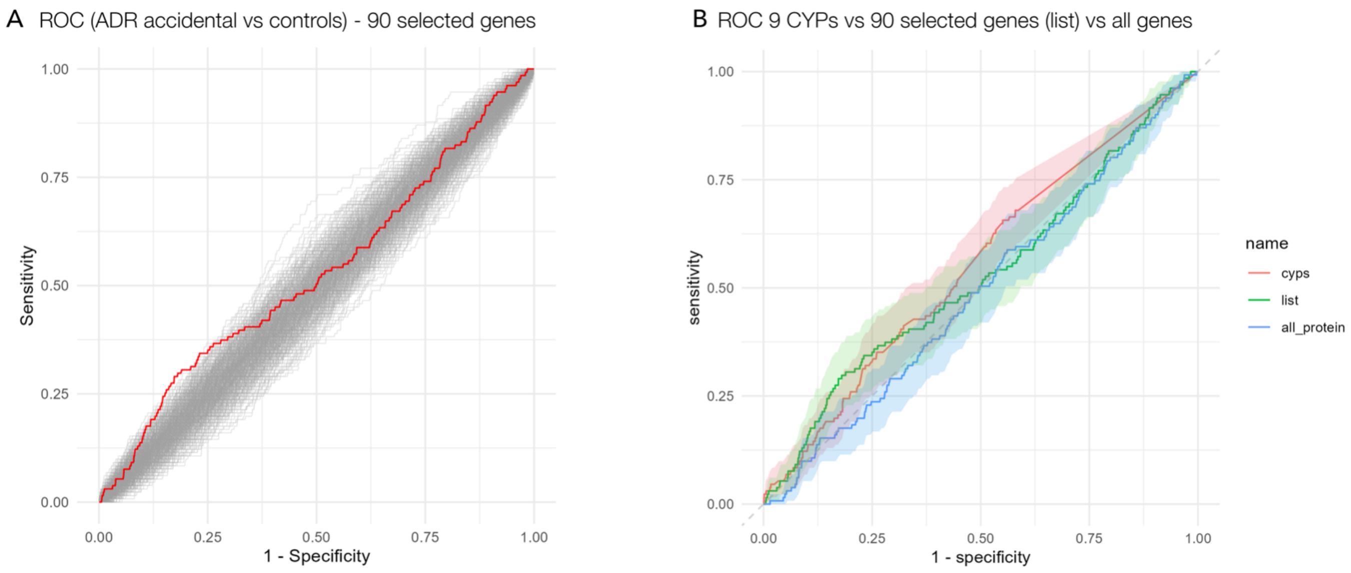


**Supplementary figure 5.** A. ROC curve of a classifier distinguishing participants of the ADR-accidental group (n = 131, includes all accidental poisonings and ADRs with therapeutic doses) versus all controls (n = 199691) showing classification performance of the compound PharmGScore based on 90 genes relevant to antidepressant responses (red line). For comparison, 1000 ROCs of sets of 90 randomly drawn protein-coding genes are shown (grey lines). The AUC of the classifier based on the list of 90 genes is 0.53 and empirical p-value of this model is = 0.121, B. Comparison of three classifiers based on a compound PharmGScores: pink line - 9 CYP genes (cyps); green line - selected 90 genes (list); blue line - all protein-coding genes (all_protein). Shaded areas represent 95% Confidence Intervals
